# COVID-19 Outpatient Screening: a Prediction Score for Adverse Events

**DOI:** 10.1101/2020.06.17.20134262

**Authors:** Haoqi Sun, Aayushee Jain, Michael J. Leone, Haitham S. Alabsi, Laura Brenner, Elissa Ye, Wendong Ge, Yu-Ping Shao, Christine Boutros, Ruopeng Wang, Ryan Tesh, Colin Magdamo, Sarah I. Collens, Wolfgang Ganglberger, Ingrid V. Bassett, James B. Meigs, Jayashree Kalpathy-Cramer, Matthew D. Li, Jacqueline Chu, Michael L. Dougan, Lawrence Stratton, Jonathan Rosand, Bruce Fischl, Sudeshna Das, Shibani Mukerji, Gregory K. Robbins, M. Brandon Westover

## Abstract

**Background:** We sought to develop an automatable score to predict hospitalization, critical illness, or death in patients at risk for COVID-19 presenting for urgent care during the Massachusetts outbreak.

**Methods:** Single-center study of adult outpatients seen in respiratory illness clinics (RICs) or the emergency department (ED), including development (n = 9381, March 7-May 2) and prospective (n = 2205, May 3-14) cohorts. Data was queried from Partners Enterprise Data Warehouse. Outcomes were hospitalization, critical illness or death within 7 days. We developed the COVID-19 Acuity Score (CoVA) using automatically extracted data from the electronic medical record and learning-to-rank ordinal logistic regression modeling. Calibration was assessed using predicted-to-observed ratio (E/O). Discrimination was assessed by C-statistics (AUC).

**Results:** In the development cohort, 27.3%, 7.2%, and 1.1% of patients experienced hospitalization, critical illness, or death, respectively; and in the prospective cohort, 26.1%, 6.3%, and 0.5%. CoVA showed excellent performance in the development cohort (concurrent validation) for hospitalization (E/O: 1.00, AUC: 0.80); for critical illness (E/O: 1.00, AUC: 0.82); and for death (E/O: 1.00, AUC: 0.87). Performance in the prospective cohort (prospective validation) was similar for hospitalization (E/O: 1.01, AUC: 0.76); for critical illness (E/O 1.03, AUC: 0.79); and for death (E/O: 1.63, AUC=0.93). Among 30 predictors, the top five were age, diastolic blood pressure, blood oxygen saturation, COVID-19 testing status, and respiratory rate.

**Conclusions:** CoVA is a prospectively validated automatable score to assessing risk for adverse outcomes related to COVID-19 infection in the outpatient setting.

## INTRODUCTION

The COVID-19 pandemic has presented unparalleled challenges for healthcare systems around the world[1–8]. The SARS-CoV-2 virus first appeared in Wuhan, China in December 2019. The first US case was confirmed on January 20[9], followed by exponential spread within the US[10]. By the end of April, Massachusetts was the third hardest hit state, trailing only New York and New Jersey[11]. Within Massachusetts, Boston and Chelsea were epicenters for the spread of COVID-19.

In anticipation of the surge of COVID-19 patients and to help limit viral spread, Massachusetts General Hospital (MGH) closed most outpatient and urgent care clinics and set up new Respiratory Illness Clinics (RICs) on March 7, 2020 to manage patients with symptoms of a respiratory infection. These clinics were staffed by clinicians and nurses reassigned from other areas, most with little urgent care experience. COVID-19 suspected cases were also screened in the Emergency Department (ED). Due to high case volumes and precautions for staff and patients, visit duration and extent of clinical assessments for most patients were curtailed. In addition to limited clinical assessment, triage decisions were complicated by COVID-19’s biphasic clinical course. Patients who initially present with mild symptoms often later return for hospital admission, and many subsequently suffer adverse events including ICU transfer, mechanical ventilation (MV) or death. Various prediction rules have been proposed, but to our knowledge few have been prospectively validated, and most were developed for the inpatient setting rather than outpatient screening.

To help frontline clinicians appropriately triage and plan follow-up care for patients presenting for COVID-19 screening and not requiring immediate hospitalization, we developed an outpatient screening score, COVID-19 Acuity Score (CoVA), that could be incorporated into an electronic medical record (EMR) system. We used data from MGH’s newly formed RIC clinics and the emergency department. CoVA assigns acuity levels based on demographic, clinical, radiographic, and medical history variables, and provides predicted probabilities for hospital admission, ICU admission or MV, or death within 7 days.

## METHODS

### Study Population

We included data from patients seen in MGH RIC clinics and ED between March 7 and May 14. Patients were divided into two mutually exclusive cohorts, a development cohort (March 7 to May 2, 2020, n = 9381); and prospective cohort (May 3 to May 14, 2020, n = 2205).

Inclusion criteria were: (1) MGH RIC or ED visit between March 7th and May 14th, 2020; (2) age ≥ 18 years; and (3) reason for visit was cough, fever, shortness of breath, COVID-related, or potentially related (see Supplemental Table S6). Exclusion criteria were: (1) Patients seen solely for a nasopharyngeal PCR swab without a clinical assessment were excluded; and (2) Patients with visits in both development and prospective cohorts are excluded, to ensure the score is not overfit to those patients. A data flowchart is provided in Supplemental Figure S1. Note that one patient contributes only once in either cohort. In the development cohort, we randomly sampled one visit from each patient. In the prospective cohort, we chose the most recent visit from each patient.

### Primary Outcome

The primary outcome was the occurrence of an adverse event within 7 days following an outpatient medical encounter, including either hospitalization at MGH, critical illness (defined as ICU care and/or mechanical ventilation), or death. The prediction horizon was set to 7 days because this period was considered meaningful by our frontline teams for clinical decision-making (e.g., regarding whether to send a patient home or to the emergency department, or to follow up with phone calls), and because empirically, within the model development cohort most adverse events occurred within 7 days of initial presentation.

### Predictors

We selected 98 variables that were routinely available in the outpatient setting during the COVID-19 pandemic to serve as candidate predictors. These included demographic variables: age, gender, tobacco use history, most recent body mass index (BMI) within the preceding month (represented as binary variables designated as high BMI, >35kg/m^2^, and low BMI, <18.5kg/m^2^), the most recent vital signs (blood pressure, respiratory rate, heart rate, temperature, blood oxygen saturation level (SpO2)) within the preceding 3 days; COVID testing status (See Supplemental Section “Predictor encoding” for encoding details); specific symptoms suggestive of COVID-19 (anosmia, dysgeusia based on ICD-10 codes), and pre-existing medical diagnoses, coded as present or absent based on groups of billing codes (ICD-10 codes; Supplemental Table S3) in the EMR. To be coded as present, the diagnostic code had to be recorded on or before the day of presentation. The Weighted Charlson Comorbidity Index (CCI) was computed based on groups of ICDs using the “wscore” from the R package “comorbidity”[14]. We omitted race and ethnicity as predictors because (1) we have found that these variables were often unavailable or inaccurately recorded, especially ethnicity; and (2) we wanted to limit the influence of local demographic patterns on the score, which might limit the ability to generalize to external populations.

For patients who underwent chest X-ray (CXR) imaging during these encounters, we identified groups of common findings based on radiology reports that tend to indicate adverse events in patients with COVID-19. These groups were identified in two steps. First, we manually reviewed 50 CXR reports and liberally extracted key words, phrases, and word patterns describing abnormal findings. Next, a pulmonary and critical care medicine physician (LB) categorized these phrases into groups. Five groups were identified: multifocal, patterns typical for COVID-19 (pneumonia, bronchopneumonia, Acute Respiratory Distress Syndrome (ARDS)), patchy consolidation, peripheral or interstitial opacity, or hazy or airspace opacities. The phrases and groupings are shown in Supplemental Table S4. CXR predictors were coded as present, not present or unavailable.

### Data Preprocessing and Selection of Predictor Variables

We treated predictors outside of physiologically plausible ranges as unavailable (see Supplemental Table S5). Unavailable values were imputed using K-nearest neighbors (KNN)[15], where the value of K was determined by minimizing the imputation error on a subset without unavailable data by randomly masking variables according to the pattern of unavailability in the overall data. Note that we did not impute CXR predictors or COVID status for patients who did not have either of them available. Instead, for these we coded unavailability explicitly (described above).

Predictors were standardized to zero mean and unit standard deviation using z-score transformation (Supplemental Table S1). Predictors were selected for inclusion in the CoVA model in two stages. First, we used ANOVA statistics to identify predictors associated with hospitalization, ICU care or MV, or death. At 0.05 significance level, we identified 65 predictors to carry forward into the model fitting procedure (p-values shown in Supplemental Table S7). Second, we used least absolute shrinkage and selection (LASSO) regression during the model fitting procedure to select a reduced subset of highly predictive variables.

### Model Development

We assigned an ordinal scale to adverse events, including no event, hospitalized, ICU care and/or MV, and death. We used an implementation of ordinal regression: pairwise learning to rank (LTR) with LASSO penalized logistic regression[16,17]. Training involves learning to predict which of a pair of patients will have a worse outcome. On biological grounds, and to address co-linearity among predictors, we constrained the model optimization to allow only non-negative coefficients for CXR predictors, medical comorbidities, CCI, and history of current or past tobacco use; and unconstrained for other predictors. Model training and preliminary evaluation of model performance was performed using the development cohort, using nested 5-fold cross validation (CV) (Supplemental Materials and Supplementary Figure S3). The final model provides acuity scores between 0 to 100 and predicted probabilities for hospitalization and for critical illness or death within 7 days. The final model was tested on the prospective validation cohort. Performance is reported both for the development cohort (results from cross validation), and for the prospective cohort.

### Statistical Analysis of CoVA Predictive Performance

We summarized the distribution of cohort characteristics and adverse events using event counts and proportions for categorical predictors, and mean and standard deviation for continuous variables. For model calibration, an expected-to-observed ratio (E/O)[18] of 1.0 indicates that the number of expected events equals the number of observed events; and calibration slope (CS) is defined as the linear correlation between the observed O and expected probabilities E, where the expected (predicted) values (E) are binned into quintiles.

We calculated the area under the Receiver Operator Characteristic (ROC) curve (AUC, also called the C-statistic) to quantify how well CoVA scores discriminate between individuals who within 7 days were vs. not hospitalized, and between those who did vs. not experience critical illness (ICU care, MV, or death). We considered an AUC between 0.50 and 0.55 to be poor; 0.55 and 0.65, moderate; 0.65 and 0.75, acceptable; and >0.75, excellent. We also calculated specificity, positive predictive value (PPV), and negative predictive values (NPV) at the 90% sensitivity level.

We also looked at the time course of adverse events as a function of CoVA score over 4 weeks (28 days) following initial presentation to the RIC or ED to understand the properties of CoVA score.

## RESULTS

### Cohort Characteristics

From March 7 to May 2, 2020, 9381 patients met inclusion criteria and were included in the development cohort. The average age was 51 years old with 49% being female. Among these, 3344 (35.6%) had adverse events within 7 days of presentation: 2562 (27.3%) were hospitalized, 679 (7.2%) received ICU care and/or were mechanically ventilated, and 103 (1.1%) died.

From May 3-14, 2205 additional patients met inclusion criteria and were included in the prospective cohort. The average age was 53 years old with 49% being female. Among these, 726 (32.9%) had adverse events: 575 (26.1%) were hospitalized, 139 (6.3%) received ICU care and/or were mechanically ventilated, and 12 (0.5%) died.

Cohort characteristics are summarized in Table 1. Compared to the model development period, during the prospective validation period there were modest increases in the proportion of patients with CXRs (prospective 52.3% vs. development 41.0%), the proportion of outpatient evaluations performed in RIC clinics (prospective 26.5% vs. development 21.4%), and in testing rates for COVID-19 (prospective 78.8% vs. development 59.5%). Several other small but likely clinically insignificant differences between cohorts also reached statistical significance, due to the large cohort sizes. COVID-19 infections and clinical adverse events by age and decade of life are shown in Supplemental Figure S2.

**Table 1.**
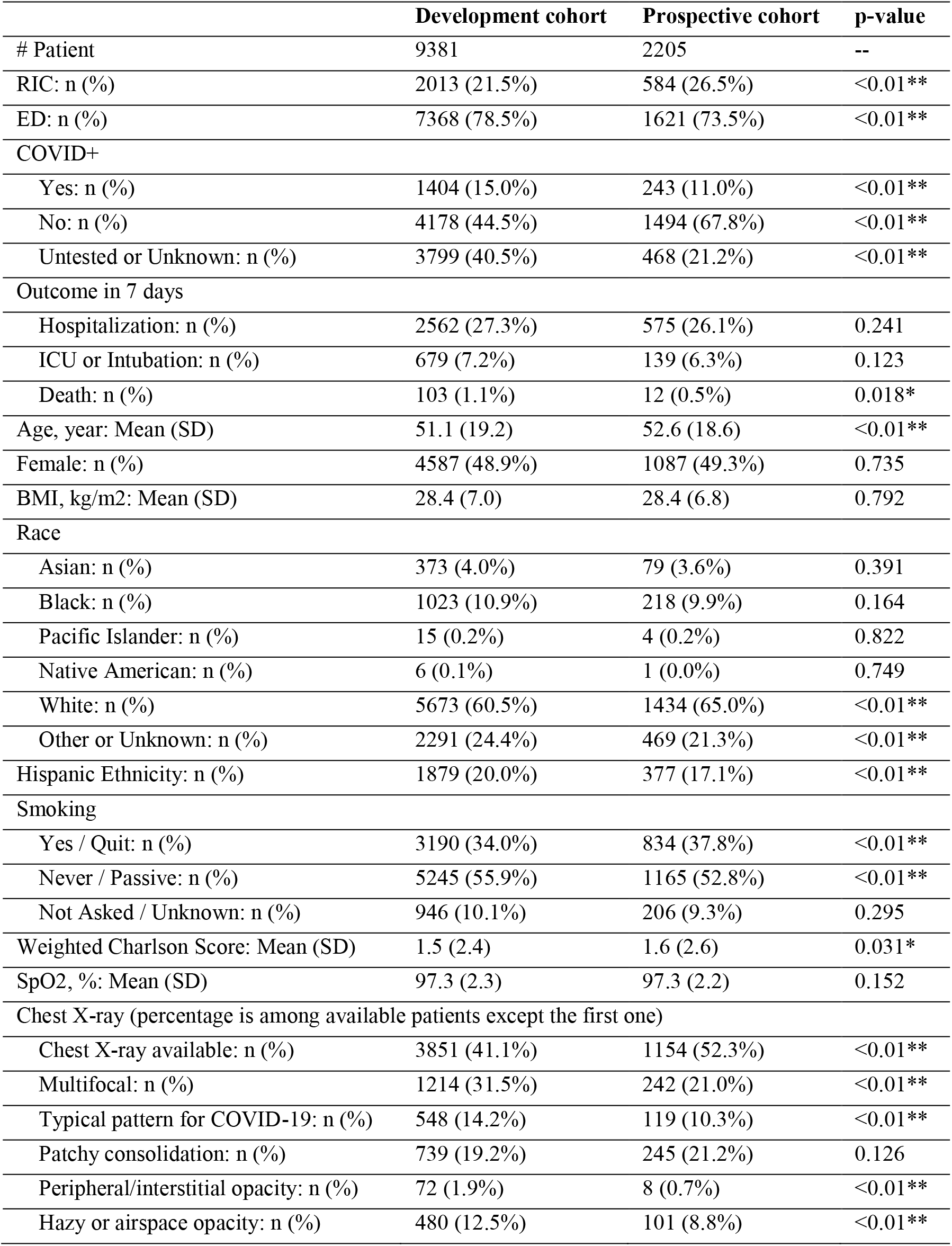
Characteristics of Study Participants. Abbreviations: RIC = respiratory illness clinic, ED = emergency department, ICU = intensive care unit, BMI = body mass index, HR = heart rate, SpO2 = oxygen saturation; CCI = Charlson Comorbidity Index.

### Predictive Performance

CoVA showed excellent calibration and discrimination in the development cohort for hospitalization (E/O: 1.00 [0.98, 1.02], CS: 0.99 [0.98, 0.99], AUC: 0.80 [0.79, 0.81]); for critical illness (E/O: 1.00 [0.93, 1.06], CS: 0.98 [0.96, 0.99], AUC: 0.82 [0.80, 0.83]); and for death (E/O: 1.00 [0.84, 1.21], CS: not calculated due to small sample size; AUC: 0.87 [0.83, 0.91]). Performance generalized to the prospective validation cohort, with similar results for hospitalization (E/O: 1.01 [0.96, 1.07], CS: 0.99 [0.98, 1.00], AUC 0.76 [0.73, 0.78]); for critical illness (E/O: 1.03 [0.89, 1.20], CS: 0.98 [0.94, 1.00], AUC: 0.79 [0.75, 0.82]); and for death (E/O: 1.63 [1.03, 3.25], CS: not calculated due to small sample size; AUC: 0.93 [0.86, 0.98]). Additional performance metrics are reported in Table 2.

**Table 2.** Prediction performance. Abbreviations: AUROC = area under the receiver operating characteristic curve, PPV = positive pressure ventilation, NPV = negative predictive value.

| Adverse event | Hospitalization, ICU, Intubation, or death | ICU, Intubation, or death | Death |
| --- | --- | --- | --- |
| **Concurrent validation (based on the development cohort but cross-validated)** |  |  |  |
| Number of patients (n, %) | 3344 (35.6%) | 782 (8.3%) | 103 (1.1%) |
| Calibration (E/O) | 1.00 [0.98, 1.02] | 1.00 [0.93, 1.06] | 1.00 [0.84, 1.21] |
| Calibration slope (CS) | 0.99 [0.98, 0.99] | 0.97 [0.96, 0.99] | n.c |
| AUC | 0.80 [0.80, 0.81] | 0.82 [0.80, 0.83] | 0.87 [0.83, 0.91] |
| Spec. at 90% Sens. | 0.56 [0.53, 0.59] | 0.44 [0.41, 0.48] | 0.35 [0.24, 0.45] |
| PPV at 90% Sens. | 0.47 [0.45, 0.49] | 0.16 [0.14, 0.17] | 0.03 [0.02, 0.04] |
| NPV at 90% Sens. | 0.89 [0.88, 0.90] | 0.98 [0.98, 0.99] | 1.00 [1.00, 1.00] |
| **Prospective validation** |  |  |  |
| Number of patients (n, %) | 726 (32.9%) | 151 (6.8%) | 12 (0.54%) |
| Calibration (E/O) | 1.01 [0.96, 1.07] | 1.03 [0.89, 1.20] | 1.63 [1.03, 3.25] |
| Calibration slope (CS) | 0.99 [0.98, 1.00] | 0.98 [0.96, 0.99] | n.c |
| AUC | 0.76 [0.73, 0.78] | 0.79 [0.75, 0.82] | 0.93 [0.86, 0.98] |
| Spec. at 90% Sens. | 0.66 [0.62, 0.70] | 0.53 [0.43, 0.63] | 0.29 [0.01, 0.31] |
| PPV at 90% Sens. | 0.40 [0.38, 0.43] | 0.11 [0.09, 0.14] | 0.017 [0.01, 0.22] |
| NPV at 90% Sens. | 0.87 [0.86, 0.89] | 0.98 [0.98, 0.99] | 1.00 [1.00, 1.00] |
| + 95% confidence interval<br>PPV = positive predictive value, NPV = negative predictive value, Spec = specificity, Sens = sensitivity, ICU = intensive care unit, E/O = ratio of Expected to number of Observed adverse events, n.c. = not calculated due to small sample size. |  |  |  |

### Properties of COVA Score

The proportion of patients with adverse events at 7 days increases with higher CoVA scores, rising from 18% with CoVA scores in the 0-20 range, to 88% for those with scores in the 80-100 range. The proportion underwent critical illness or death also rises, from 2% for scores between 0-20, to 32% with scores between 80-100 (Figure 1A).

**Figure 1.**
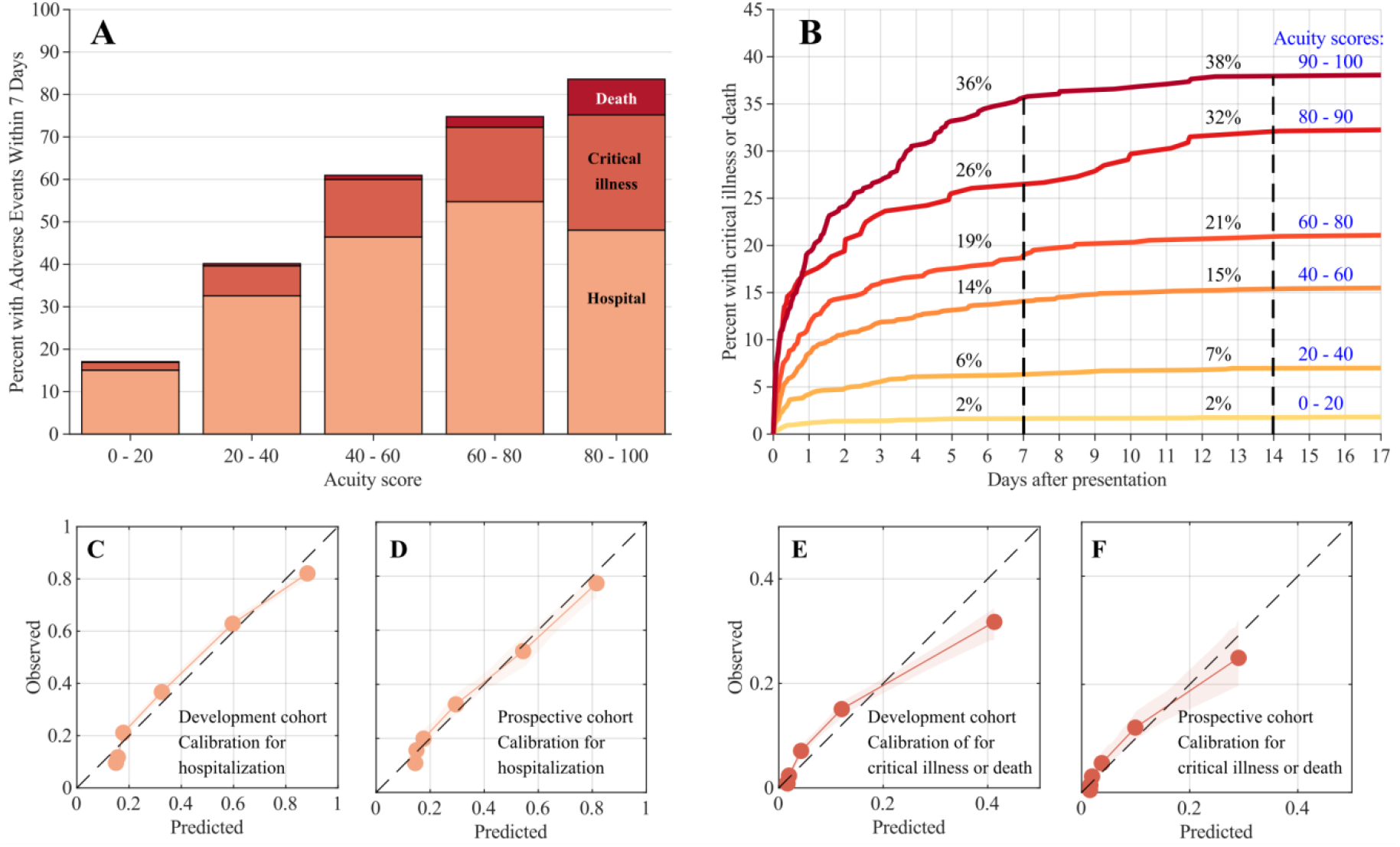
(A) Distributions of adverse events (AE) within 7 days after initial outpatient evaluation in the RIC/ED, binned by acuity score. Colors represent hospitalization, ICU/MV, or death. (B) Cumulative incidence of critical illness or death up to 17 days following initial evaluation, based on initial acuity score. Curves are computed based on cross validation in the development cohort (C,D,E,F) Calibration curves: predicted probability of adverse events vs. observed rate of adverse events. C (development cohort) and D (prospective validation cohort) show calibration for predicting hospitalization; E (development cohort) and F (prospective validation cohort) show calibration for predictions of critical illness or death. Overall calibration (E/O) and calibration slopes (CS) are reported in Table 2.

We investigated the time course of adverse events as a function of CoVA score over 4 weeks (28 days) following initial presentation to the RIC or ED. We limited this investigation to the development cohort, because 28 days have not passed for the prospective validation cohort at the time of writing. By 28 days, 3399 (36%) of patients experienced hospitalization, critical illness, or death. Of these, 3214 (95%) occurred within 1 day, and 3344 (98%) had occurred by 7 days. Critical illness or death occurred in 861 (9%) of patients within 28 days of presentation. Of these, 537 (62%) occurred within 1 day, and 782 (91%) occurred within 7 days. These numbers support our choice of 7 days as a clinically meaningful event prediction horizon. Curves for the cumulative incidence of adverse events over time for different levels of CoVA scores are shown in Figure 1B.

### Predictors of Adverse Events

Thirty predictors were selected by the data-driven model training procedure (Table 3). All but four predictors increase the predicted probability of adverse events when present. SpO2, diastolic and systolic blood pressure and low BMI inversely correlated with the probability of adverse events. Predictors from CXR reports included in the model were multifocal patterns (diffuse opacities, ground glass) and patterns typical for COVID.

**Table 3.** Coefficients of the CoVAS model.

| ID | Predictor | coefficient | ID | Predictor | coefficient |
| --- | --- | --- | --- | --- | --- |
| 1 | Age | 0.7353 | 16 | Renal cancer* | 0.0681 |
| 2 | Diastolic blood pressure | -0.4724 | 17 | Pancreatitis* | 0.0608 |
| 3 | SpO2 | -0.3776 | 18 | Cystic fibrosis* | 0.0492 |
| 4 | Ever COVID+ up to event | 0.2750 | 19 | Cardiac Arrest* | 0.0491 |
| 5 | Respiratory Rate | 0.2746 | 20 | Seizure disorder | 0.0437 |
| 6 | Acute ischemic stroke* | 0.1746 | 21 | Amyolateral sclerosis* | 0.0405 |
| 7 | CXR: Multifocal | 0.1293 | 22 | Metabolic acidosis* | 0.0385 |
| 8 | Heart rate | 0.1215 | 23 | Myasthenia gravis* | 0.0374 |
| 9 | Body temperature | 0.1206 | 24 | Pneumothorax* | 0.0300 |
| 10 | Systolic blood pressure | -0.1151 | 25 | Spinal muscular atrophy* | 0.0241 |
| 11 | Weighted Charlson Score | 0.1142 | 26 | Pericarditis* | 0.0144 |
| 12 | Intracranial hemorrhage* | 0.1087 | 27 | High BMI (>35kg/m<sup>2</sup>) | 0.0028 |
| 13 | Subarachnoid hemorrhage* | 0.0919 | 28 | CXR: Typical for COVID | 0.0001 |
| 14 | Male sex | 0.0808 | 29 | Low BMI (<18.5kg/m<sup>2</sup>) | -0.0001 |
| 15 | Hematologic malignancy* | 0.0765 | 30 | ARDS* | 0.0001 |
| **Legend:** Abbreviations: CXR = chest X-ray; ARDS = Acute Respiratory Distress Syndrome; BMI = Body Mass Index. Diagnoses are based on past medical history, and are coded as present (e.g. Pneumothorax = 1) if recorded in the electronic medical record at any time before the date of presentation for COVID-19 screening. Disorders marked by ‘*’ refer to pre-existing conditions documented in the electronic medical record. |  |  |  |  |  |

### Effect of Chest X-Ray Findings and COVID Status on the Probability of Adverse Events

CXRs and testing for COVID-19 were not universal, and testing rates evolved over time. One potential use of CoVA is to help determine whether to perform these tests. We therefore investigated the impact of CXR findings on the predicted probability of critical illness or death. As shown in Supplemental Figure S4, positive CXR findings are most informative when the pre-CXR probability is 30% (critical illness or death), in which case they increase the predicted probability of an adverse event by 4%. Negative CXR findings are maximally informative when the pretest probability is near 34% (critical illness or death), in which case they decrease the probability by 4%.

We also examined the effect of COVID-19 testing results on the predicted probability of adverse events (critical illness or death). The largest effect of a positive test result occurs when pretest probability is 28%, in which case the post-test probability increases by 8%. A negative result has the largest impact when pretest probability is 36%, in which case the posttest probability decreases by 8%.

## DISCUSSION

Identifying outpatients presenting for COVID-19 screening at high risk or low risk for adverse events is important for medical decisions regarding testing, hospitalization, and follow-up. We developed and prospectively validated the COVID-19 Acuity Score (CoVA) as an automatable outpatient screening score that can be implemented in the EMR. The model exhibits excellent calibration, discrimination, and negative predictive value both in concurrent validation (n = 9381, March 7-May 2) and in large-scale prospective validation (n = 2205, May 3-14). While several COVID-19 risk prediction models have been proposed for the inpatient setting, CoVA fills an unmet need for a prospectively validated risk designed for outpatient screening.

Several predictors selected into the model have been identified in prior studies, including advanced age[4,5,9,19]; pre-existing pulmonary[4,5], kidney[4], and cardiovascular disease[4,9,20]; obesity[21]; and increased respiratory or heart rate or hypoxia[4]. We found that other pre-existing medical conditions also increased risk for adverse outcomes, including hematologic malignancy, cancer and pancreatitis. Hypertension and diabetes mellitus did not emerge as predictors in the CoVA model, despite being identified in prior studies[4, 5, 9]. These comorbidities are correlated with outcomes in univariate analysis (Supplemental Table S7) and correlation analysis shows that both are strongly associated with older age, higher CCI, high BMI, and other comorbidities already selected as predictors, helping to explain their not being selected into the model (Supplemental Figure S5). HIV/AIDS was not significantly associated with the outcome, but this may have been due to low numbers (85/9381<1%, less than 1% of developmental cohort).

Several studies have documented neurological manifestation of COVID-19[22–24]. Nevertheless, we were surprised to find a variety of neurological diseases surfaced as robust predictors of adverse outcomes in COVID-19 infection, including ischemic stroke, intracranial hemorrhage, subarachnoid hemorrhage, epilepsy, amyotrophic lateral sclerosis, myasthenia gravis, and spinal muscular atrophy. It is unclear if these neurological diseases are merely markers of health or if the worsened outcomes are due to the interaction of COVID-19 with neurological disorders that amplifies the pathology.

Prior work on predicting outcomes in COVID-19 patients is summarized in Supplemental Table S2. Like our study, most have attempted to predict critical illness or death. However, most are based on small cohorts (median: n = 189, range n = 26 to 577); focus on inpatients; and utilize laboratory values, which were rarely available for our outpatient cohort. Only 3 included prospective or external validation. By contrast, CoVA is designed for the outpatient setting. In this setting, the availability of COVID-19 test results are variably available (for 60% in the development and 80% in the prospective cohort) and other laboratory results were rarely available. To ensure generalizability, we used a large development cohort of 9381 patients, and trained the model using a rigorous approach. We ensured clinical interpretability by utilizing a linear model with positivity constraints on predictors expected to increase risk. Finally, we validated our model on a large (n = 2205) prospectively collected patient cohort, providing an unbiased assessment of model generalizability.

Strengths of this study include its large sample size, careful EMR phenotyping, rigorous statistical approach, and the feature that all variables required by the model are available and automatically extractable within most electronic health record systems. The study also has limitations. First, our study is from a single center, with patient demographics specific to MGH, Boston. Nevertheless, important biological parameters in the predictive model are universal and increase the likelihood that the model will generalize. Second, although RIC clinics were established for patients with suspicion for COVID-19, patients seen in the ED were seen for a variety of reasons. Nevertheless COVID-19 was and for now remains a universal concern for patients seen in the ED, and we excluded patients prior to the onset of COVID-19 in Boston (March 7, 2020), therefore our model is relevant for screening during times of high alert for COVID-19. Finally, we did not include lab test values, since they were typically not available. Among the few studies of inpatients, inclusion of lab tests appears to improve model performance.

In conclusion, the COVID-19 Acuity Score (CoVA) represents a well-calibrated, discriminative, prospectively validated, and interpretable score for assessing the risk for adverse events for outpatients presenting with possible COVID-19 infection.

## Data Availability

The data is available upon request from the corresponding author if reasonable.

## Acknowledgments

The authors thank all of the physicians and nurses of Massachusetts General Hospital for their exemplary patient care. The authors also gratefully acknowledge all of the physicians of the Respiratory Infection Clinics in the MassGeneral Brigham hospital system, and the MGH Emergency Departments for their constructive feedback on this work.

